# Interventions to promote completion of human papilloma virus vaccination course in high-risk groups attending a London sexual health clinic

**DOI:** 10.1101/2025.11.12.25340109

**Authors:** George Budden, Sophie Monouchous, Samantha Stancu

**Affiliations:** Homerton Healthcare NHS Foundation Trust

## Abstract

**Objectives:** Men who have sex with men (MSM) have a 30-fold higher risk of anal cancer compared to the general population and the Human Papilloma Virus (HPV) vaccine can reduce this risk.

Completion rates of the vaccine course remain low. The aim of this project was to improve vaccine course completion rates in high-risk groups.

**Methods:** Patients not returning for the second dose of the vaccine over a 12-month period were sent text message reminders. After four weeks, analysis was done on those who returned. Other interventions to increase vaccination uptake comprised a patient survey, updating pertinent information on the clinic website and displaying posters in clinic rooms.

**Results:** The vaccine course completion rate increased from 37% to 39%. Our patient survey showed 87.5% of responders would opt for text message reminders about subsequent vaccine doses.

**Conclusions:** Human Papilloma Virus vaccine course completion rates remain below the NHS England target. A user-friendly and time-efficient recall system was devised in order to enhance vaccine course completion. Future strategies could include creating a vaccine passport, providing more in-depth counselling on the importance of HPV vaccination and creating an opportunistic walk-in appointment service for vaccination at sexual health clinics.

## Introduction

Human Papilloma Virus (HPV) is the leading cause of anal and cervical cancer worldwide. It is the causative agent of numerous other types of cancers (penile, vaginal, oropharynx) in addition to genital warts (1). Although an effective vaccine already exists that can reduce the risk of infection and its sequelae, vaccine completion rates amongst certain high-risk groups such as gay, bisexual and other men who have sex with men (GBMSM) as well as those living with Human Immunodeficiency Virus (HIV) remains below the national target of 55% (2). These populations have a substantially higher risk of anal cancer, with GBMSM having an approximate 30-fold increased risk compared to the general population (3,4). Hence, strategies to increase HPV vaccination rates are of upmost importance.

The United Kingdom’s HPV vaccination programme was initially implemented for 12-13 year-old girls and achieved a 90% reduction in prevalence of HPV strains 16 and 18 (5). Herd immunity effects were seen in boys too. However, a large proportion of the remaining population was left unvaccinated and unprotected (6). Since 2018, this programme was extended to a national two-dose course for GBMSM aged 25 to 45, using the 9-valent Gardasil ™ HPV vaccine (1). Those younger than 25 only require a single dose and those living with HIV require three doses. To improve the uptake of this programme, the National Institute for Health and Care Excellence (NICE) guidelines suggest sending reminders to those eligible who have not returned to emphasise the importance of completing the course (7).

This quality improvement project aimed to promote the completion of HPV vaccination courses in high-risk groups at a London sexual health clinic. Interventions included sending text message reminders and promoting knowledge about HPV to both patients and staff through posters in clinic. We also conducted a patient survey to find out about how patients would prefer to receive reminders. The end-goal was creating a recall system for those receiving their first dose to increase enhance course completion rates.

## Methods

This quality improvement project took place across four clinics in the City and Hackney Borough of London.

Consecutive patient records were extracted by the Information Technologies (IT) team of patients who received their first of two doses of the HPV vaccine in a 12-month period, between 1^st^ July 2023 and 31^st^ June 2024, who had not returned for a second dose.

All patients who consented to contact via text message were sent a text message informing them they may need a further dose and instructions on how to book an appointment. The message explained they may have had subsequent doses at other clinics to prevent patients re-attending when they had already completed their vaccine course elsewhere.

A patient survey was conducted to gain insight into patient preferences on how they would want to be contacted to return for subsequent doses of vaccinations and about any potential barriers to accessing vaccine doses.

The clinic website was updated with information about vaccination schedules and posters about the vaccinations were displayed in clinics.

After a four-week period to allow patients time to return, records were extracted of patients who had received a second dose of the HPV vaccine and cross-referenced with our initial patient list to extract a list of patients who returned following text message reminder.

## Results

Five hundred and sixty-one patients received the first dose of the HPV vaccine in the 12-month timeframe. As depicted in Table 1: 97.5% were male, 54.7% were aged 25-34, 43.1% were from Hackney and 83.2% identified as homosexual.

**Table 1:**
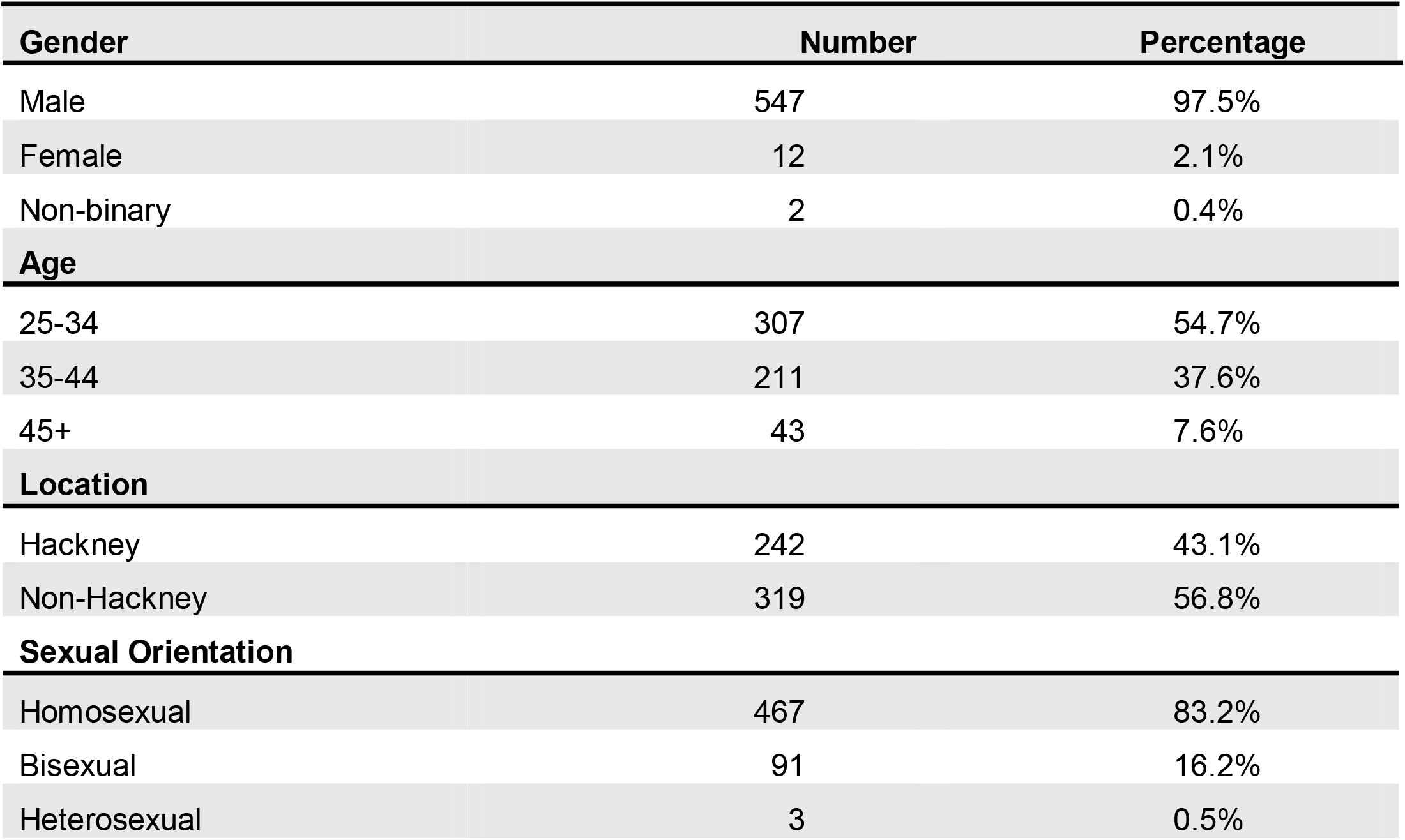
Demographics of patients who received a first dose of the vaccine in the 12-month window.

| Gender | Number | Percentage |
| --- | --- | --- |
| Male | 547 | 97.5% |
| Female | 12 | 2.1% |
| Non-binary | 2 | 0.4% |
| **Age** |  |  |
| 25-34 | 307 | 54.7% |
| 35-44 | 211 | 37.6% |
| 45+ | 43 | 7.6% |
| **Location** |  |  |
| Hackney | 242 | 43.1% |
| Non-Hackney | 319 | 56.8% |
| **Sexual Orientation** |  |  |
| Homosexual | 467 | 83.2% |
| Bisexual | 91 | 16.2% |
| Heterosexual | 3 | 0.5% |

Two hundred and twelve patients (37.7%) of patients who received their first dose returned for their second dose within the outlined timeframes without intervention. Forty-six percent of patients were from City & Hackney compared to 41% of those who did not return.

Three hundred and fifty-nine patients did not return for their second dose. After removing patients who did not consent to contact via text message, 339 patients were texted to return.

Of these 339 patients, 8 returned within the four-week timeframe to complete their vaccination course. This increased the total completed vaccination rate from 37.7% to 39.2%. Of the 8 patients who returned, the mean age was 36.6 years and 50% were from City and Hackney.

The survey revealed that 87.5% of patients would prefer a text reminder to return for subsequent doses.

When asked about barriers preventing patients from completing their vaccination course, struggling to keep track of which vaccine was needed and when, difficulty booking appointments and forgetting were the most common responses.

## Discussion

Despite patients reporting text-message reminders as the preferred method of communicating when subsequent vaccine doses are due, this intervention did not significantly increase HPV vaccine course completion. This could be attributed to the content, amount and method of sending reminders. Vollrath et al. suggest that there are mixed results with using a single method of vaccination recall alone, although it has similar effectiveness in increasing recall rates compared to using multiple methods of recall (8). In their study, phone calls were the most effective method of vaccine recall, although this is labour intensive.

The content of the text message reminders could also be an influencing factor, as sending a message of eligibility alone, may not be as effective. Choi et al. found that the content of reminders and method of communication was not found to significantly increase engagement of women receiving communications regarding their HPV care (9).

Research in a UK sexual health centre found the most common reason for not returning for subsequent doses of HPV vaccinations was patients believing they were not eligible to receive the next dose of the vaccine yet. This alludes to the importance of educating patients about vaccination schedules to encourage course completion. Other reasons included the challenge of securing an appointment. When surveyed, similarly to our results, they found the majority would prefer a text message reminder (10).

### Strengths

This project was devised as a prospective study and identified that text message reminders may not be the most effective intervention in promoting the completion of the HPV vaccination course. By surveying patients first on their preferred method of communication, we were able adopt an informed approach to our intervention of choice. Our sample size was representative of a large urban sexual health service in a UK city.

### Limitations

The main limitation of this study was the fact that patients were given a four-week timeframe to return, whereas, a six-month grace period is normally given. As a result, the true extent of those returning may have not been accurately portrayed. Due to the complexity of eligibility, the text message itself required the patient to determine whether they were eligible based on their characteristics, posing an additional barrier to recall. Patients may have attended other clinics, affecting the number of returners recorded. Our survey showed that some barriers include the inability to easily book a sexual health appointment for patients which is systemic to the NHS, however, addressing this would improve risk reduction.

In conclusion, sexual health centres are best equipped to provide HPV vaccination and encourage course completion. Along with a straightforward, user-friendly and time-efficient recall system, we propose a vaccine passport where vaccination records are kept to improve engagement with sexual health vaccinations and to provide clarity for patients. Creating a small number of walk-in appointments could also facilitate accessibility for patients wishing to complete their vaccination course. Importantly, this quality improvement project has paved the way for devising a recall system for improving completion course of other vaccines for high-risk groups, namely Hepatitides A and B, Monkeypox and the Bexsero™ (4CMenB) meningococcal B vaccine that has 30-40% cross-protection against *Neisseria gonorrhoeae* infections (11).

## Data Availability

All data produced in the present study are available in anonymised format upon reasonable request to the authors

